# The Math of Margins: A Fresh Look at Bread Loaf Sections, Unchallenged Assumptions, and Citogenesis

**DOI:** 10.1101/2022.06.25.22276453

**Authors:** Melissa M. Warne, Matthew M. Klawonn, Robert T. Brodell

## Abstract

Although there are many possible ways to treat skin cancer, most skin cancers are effectively treated by complete excision followed by standard histologic evaluation to ensure clear margins. The bread loaf technique describes a common method of processing specimens in which multiple slices of tissue are taken perpendicular to the major axis of an excision and submitted for microscopic analysis. Although sections may only be approximately four microns thick (0.000004 meters), this method is associated with high cure rates for basal cell and squamous cell carcinoma. Some authors have stated that this technique assesses less than 1% of the margins. We critically reviewed this assumption. While we confirm that the bread loaf technique often directly visualizes 1% or less of the peripheral and deep margins when considering only the width of sections compared to the entire length of an excisional specimen of the excision, much useful additional information is gained as soon as clear sections are identified towards the tips of a typical excisional specimen. For tumors that tend to grow in a nodular or spherical arrangement such as nodular basal cell carcinoma or squamous cell carcinoma of keratoacanthomatous type, we show that a variable but significant portion of the margin may be considered sampled by proxy when slice faces are clear. We highlight the importance of understanding the principles involved in tissue sectioning in order to allow clinicians to make informed decisions on behalf of patients.

## 1 Introduction

Management of skin cancer typically involves excisional surgery. Ascertaining tumor-free margins requires thoughtful sampling, staining, and grossing procedures.[1] One commonly used technique involves multiple sections taken perpendicular to the long axis of the surgical specimen. This technique has been called the “bread loaf” method because the sections taken result in sections reminiscent of the slices of sliced bread. The idea that bread loaf sectioning assesses 1% or less of surgical margins has persisted in the medical literature for more than 40 years and is cited on the internet and in textbooks [2, 3, 4, 5, 6]. In reviewing this literature, it becomes clear that authors have considered the lateral and deep margins in their calculations but typically neglect the other pertinent margins, including only 3 of 5 relevant surfaces [6, 7]. We explore the geometry of a typical excisional specimen, and provide a formula that approximates the percentage of margin analyzed when the distance between clear planes perpendicular to the major axis, length, width, and height of the specimen are known. We concede that the percentage of peripheral and deep margin *directly* analyzed is small and can be considered negligible, but argue that the value of understanding the tumor biology of various histologic sub-types of skin cancer, specifically basal cell carcinoma and keratinocytic carcinoma, and the consequence of identifying tumor free planes provides additional information that accounts for the good cure rates associated with standard processing methods[8].

Simple excision often entails removal of a fusiform specimen with a length that is approximately 3 times the width and with apical angles of 30 degrees [9]. Tissue contraction and fixation result in a specimen that is often shaped like an elliptical cylinder. The excision is deliberately centered around the visible portion of the cancer [10, 11]. Many surgeons mark out the tissue that they tend to excise with a predetermined margin of clinically normal skin [8]. When inspecting the excisional specimen, the goal is to determine whether or not the margin is *clear*, or cancer free. Bread loaf sectioning entails taking slices of tissue perpendicular to the major axis and the bases of the cylinder (parallel to the minor axis) (see Fig. 1). Traditionally [4], the percentage of margin assessed by a single slice during bread loaf sectioning is calculated as the area of the intersection of the margin (cylinder’s side) and slice thickness, divided by the total area of the margin. This is illustrated in Fig. 2

**Figure 1:**
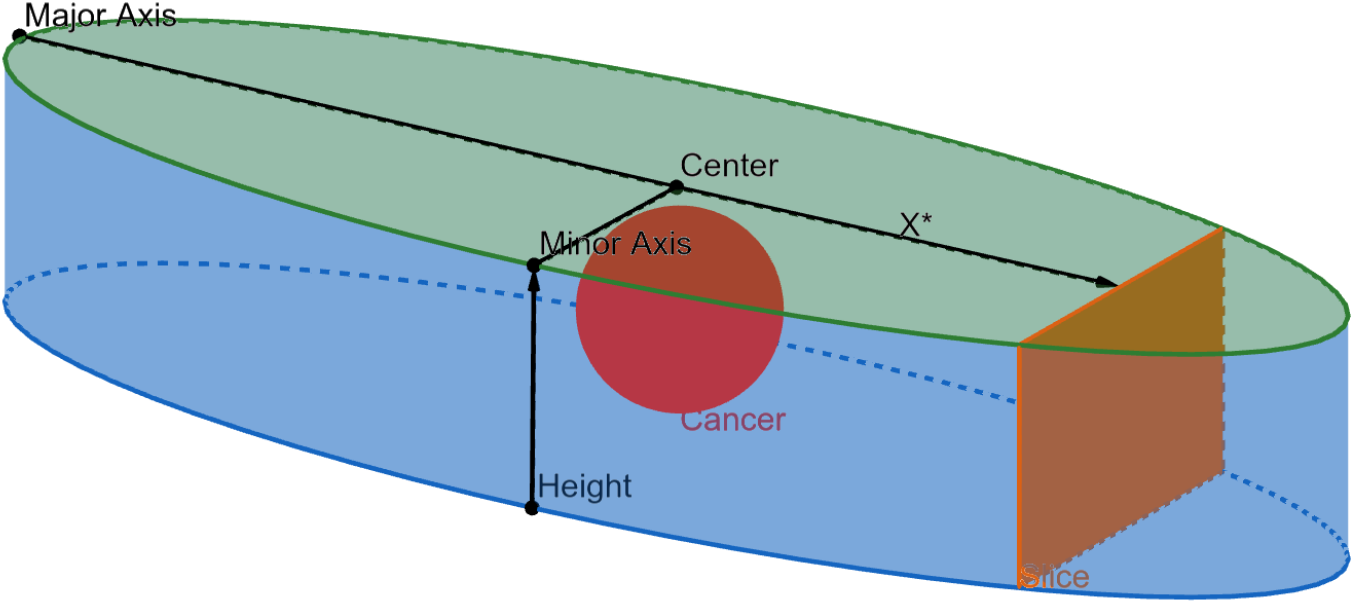
The typical shape of an excision after tissue contraction and fixation. Shows a slice/section (orange), with clear face, of distance *x*^*∗*^ from center. Cancer would typically be visible from top (green) face. The blue faces of the elliptical cylinder (side and bottom) are the margin.

**Figure 2:**
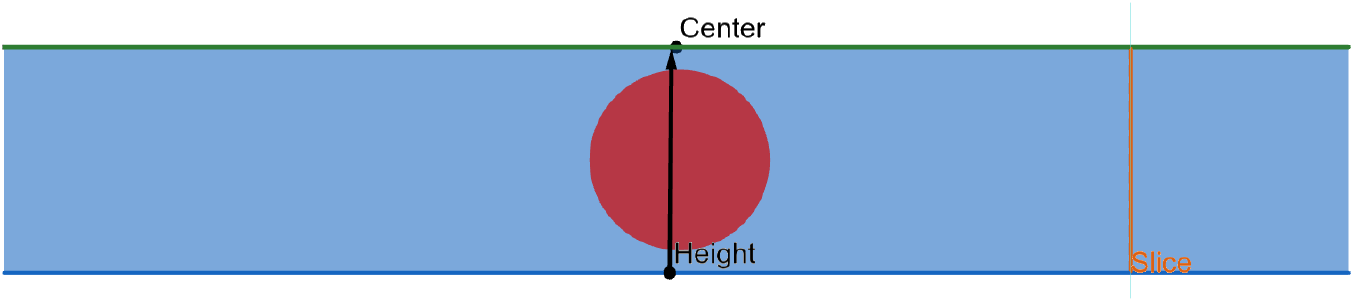
Portion of margin (orange) typically considered sampled by prior literature.

While this intersection does capture the only portion of the margin *directly* observed by bread loaf sectioning, we argue that a significantly larger portion of the margin may be considered as *observed by proxy*. The critical observation is that skin cancer does not grow and then cleave arbitrarily [10]. Rather, the neoplastic cells that comprise primary cutaneous basal cell and squamous cell carcinoma exhibit cellular adhesion and are *connected* in the topological sense of the word, as illustrated in Fig. 3. Neoplastic cells tend to migrate along a “path of least resistance” and migration paths can often be predicted by considering “energetic costs”[12]. In many cases, neoplastic cells follow interactions regulated by hydrodynamics.[12, 13] ***Therefore, in most situations, tumor does not pass through a clear section/slice unless the slice face is not clear of cancer***. This is assumption is very relevant to cancers that often have a roughly spherical shape such as a basal cell carcinoma of nodular type or a squamous cell carcinoma of keratoacanthomatous type. An exception to this could be a tumor that has been incompletely treated and is recurrent in multiple loci, but we choose not to consider this case for clarity of exposition, and acknowledge it as a limitation of our approach. There is also the possibility of microscopic disease so small that it evades detection by routine microscopic examination, but this is a risk encountered in all conventional methods of margin assessment.

**Figure 3:**
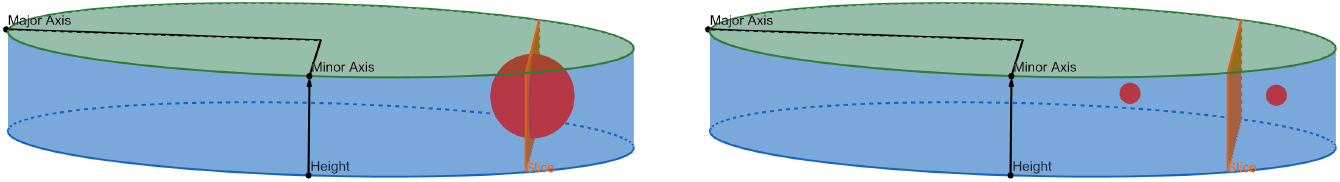
**Left**: Example of how cancer may cross a slice, the faces of which are not clear. **Right**: A hypothetically impossible example of cancer crossing a slice face by growing and cleaving.

A clear face of a slice therefore rules out the possibility that the cancer has passed the margins beyond that face for many histologic types of basal cell or squamous cell carcinoma. Further, we can calculate the percentage of the margin that has been observed by proxy when the distance between clear planes perpendicular to the major axis, length, width, and height of the specimen are known. We now give a formula for doing so.

## 2 Methods

Our computations will be broken into two parts, one corresponding to the lateral margin and one to the deep margin. We start with the lateral margin.

### 2.1 The Lateral Margin

When unrolled, the side of an elliptical cylinder is a rectangle with height *h* and width *c*, where *c* is the circumference of the ellipse. The surface area of the lateral margin, therefore, is given by the area of this rectangle, *A*_*l*_ = *c × h*. Instead of the full circumference, the margin sampled by proxy (pictured in Fig. 4) has a width equal to the arc length of the ellipse beyond the inducing clear slice. To compute the surface area of the margin sampled by proxy, we therefore need to determine this arc length (see Fig. 4). Note that these computations ignore the directly observed margin, since it is negligible at *<* 1% of the total margin.

**Figure 4:**
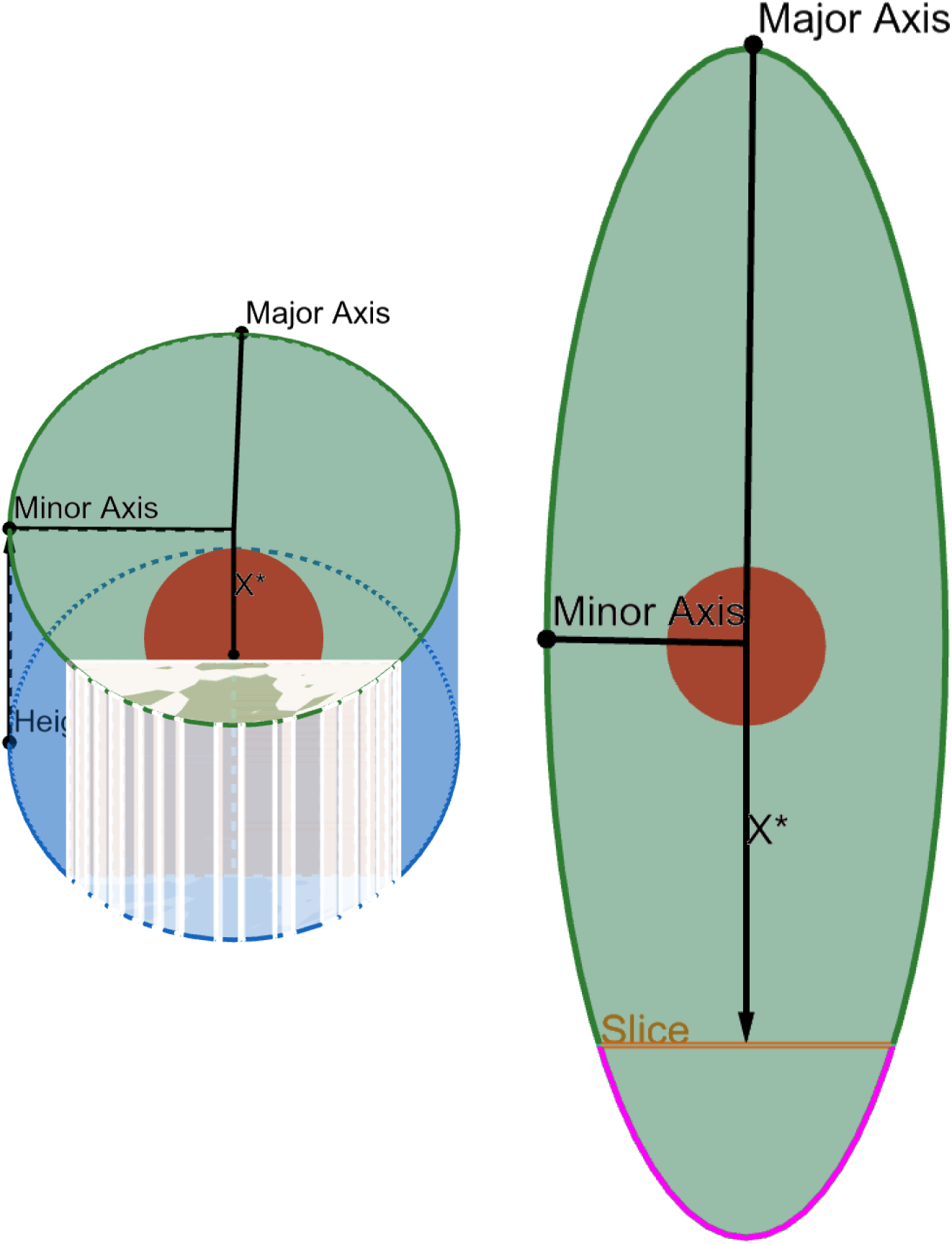
**Left**: Eq. 11 calculates the ratio of the margin in white to the whole of the margin (blue). **Right**: The area of the margin sampled by proxy depends on the the arc length pictured in pink.

Let the semi major axis length be denoted *a*, the semi minor axis *b*. The parametric equations of a location (*x, y*) on an ellipse are given by *x* = *a* cos *t, y* = *b* sin *t*. Denote by *x*^*∗*^ the distance from the center of the ellipse to the nearest clear slice on one side of the excision. In fact, *x*^*∗*^ denotes the distance from the center to the outside face of the slice, as the slice itself has a greater than zero thickness. Solving for *t*, we have *t* = arccos 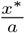. Using the arc length formula 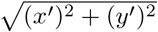 [14], and the fact that *x* = *−a* sin *t, y* = *b* cos *t*, then the arc length (denoted *l*^*∗*^) of the margin that can be considered sampled is:

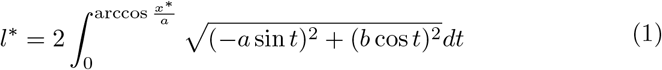

where the multiplicative factor of 2 is derived from the symmetry of the arc about the major axis.

This integral has no closed form solution, but can be computed numerically. The area of the sampled-by-proxy lateral margin 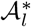 is given by multiplying *l*^*∗*^ by the height of the excision:

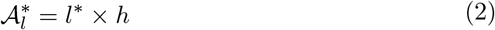

Note that this only covers one portion of the lateral margin. There will be a clear slice on the other side of the ellipse (reflected about the center along the major axis), potentially with a different distance from the focus which we will call *x*^*†*^. This yields a second arc length characterizing the margin induced by that slice. Denote by *l*^*†*^ the arc length induced by this clear slice, which can be computed using Eq. (1), swapping *x*^*∗*^ for *x*^*†*^.

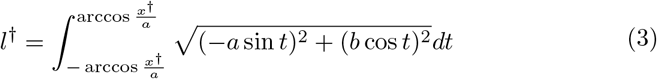

with corresponding sampled by proxy margin area:

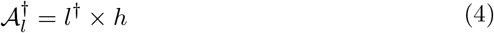

The area of the whole lateral margin is equal to the circumference of the ellipse times the height,

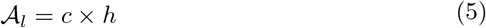

The formula for circumference *c* is 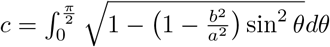 (unfortunately there is no analytical solution for the circumference of an ellipse).

With the lateral margins sampled by proxy given in Eq. (1) and (3), and the total lateral margin given in (5), we can turn our attention to calculating the deep margins sampled by proxy and the total deep margin area.

### 2.2 The Deep Margin

A clear slice, when viewed as a chord of the elliptical bottom face of the excision, defines a segment, i.e. the part of the ellipse between the slice and the induced arc. Our goal is to find the area of this segment. Rather than relying on a general formula for the area of a segment of an ellipse, we take advantage of the fact that the slice is parallel to the minor axis of the ellipse. The area of the upper half of the segment area, i.e. the half above the major axis, is given by the area under the curve of the ellipse. This is an integral of the height *y* of the curve taken over the interval [*x*^*∗*^, *a*]. To obtain the area of the segment, we double this integral (to account for the area beneath the major axis). Given an ellipse defined as 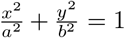. and the distance *x*^*∗*^, the area of the deep margin sampled by proxy is 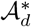.

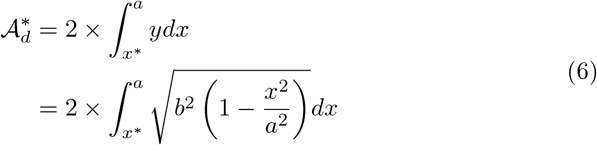

The derivation of the sampled by proxy deep margin on the other side of the excision, 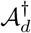, is exactly analogous, replacing *x*^*∗*^ with *x*^*†*^:

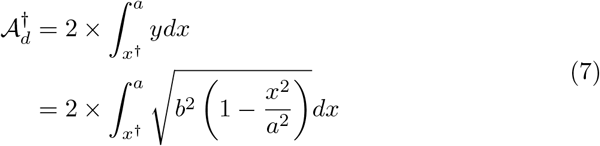

The whole area of the deep margin is simply the area of the excision’s bottom face:

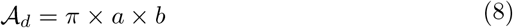

#### 2.2.1 Aside: Assumptions for Calculating the Sampled Deep Margin

Note that in assuming the deep margin can be sampled by proxy, we are relying on a tumor’s resistance to growing beneath the excisional area. Although there may be rare exceptions, neoplastic cells tend to grow along tissue planes and respect boundaries.[12, 13]. Basal cell carcinomas are now considered to be a “monoclonal proliferation of unicellular origin” in which neoplastic cells grow in contact with one another [15]. In other words, we assume that the tumor does not cross the deep margin before the clear slice, only to re-emerge through the deep margin after the clear slice. In practice, when a tumor is detected close to the examined surgical margin, conscientious pathologists will request additional sections in the vicinity of the area where tumor approximates the margin to help exclude this possibility. Some pathologists will not report a margin as “clear” if tumor is close to the margin but instead will report a distance between the tumor and the examined inked margin. Sections are sometimes taken as often as every 2-3 mm intervals [7]. Sections must not have gaps, should be cut full face, and have ink at the peripheral and deep margins to confirm that they represent true surgical margins. Unfortunately, there is no uniform standard for how frequently sections are taken transverse to the long axis and the frequency at which sections/slices are taken can vary from lab to lab.

### 2.3 Calculating Total Percentage of Margin Sampled by Proxy

Taking stock, we have now defined the lateral margins sampled by proxy, the deep margins sampled by proxy, and the total area of the lateral and deep margins. The area *A*_*s*_ sampled by proxy is the sum of the two lateral submargins and two deep submargins, given in Eqs. (2), (4), (6), and (7):

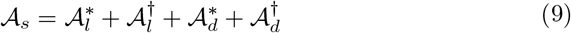

The total area of the margin is the sum of the lateral margin and deep margin, given in Eqs. (5) and (8):

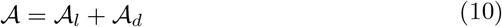

And finally, the percentage *P* of the total margin sampled by proxy can be computed as:

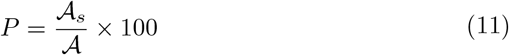

## 3 Example Calculations

To show how the percentage sampled by proxy varies given different clearance distances from the center, we give calculations of two extremes. Both of these hypothetical situations are presented as examples to show the marked variability of the bread loaf technique. The first example we will consider is for a tiny tumor at the very center of the excisional specimen that is immediately and completely removed by a single 4 micron section. The other involves a tumor that is not removed until sections taken 2 mm from the tips at each end of the specimen are found to be clear. We chose a distance of 2 mm from the tips arbitrarily with the knowledge that many histology technicians may have difficulty handling specimens smaller than this size and achieve full face sections. All integrations were performed using WolframAlpha [16].

### 3.1 Example One: Slice at Center

If we assess a hypothetical situation in which a single four micron section centered in the sample completely extirpates a tumor in an excisional specimen with a major axis length of three cm (semi-major length 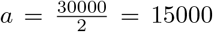 microns), a minor axis length of one cm (semi-minor length 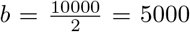 microns), and a height of one cm, the percentage of margin assessed is computed as follows. We set 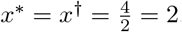. Using Eq. (1) with the appropriate values, we have

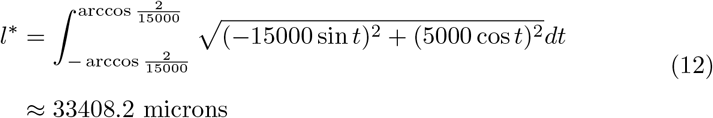

We compute the lateral margin values first. Given that *x*^*∗*^ = *x*^*†*^, we have 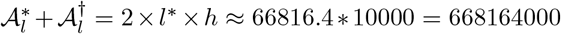 microns squared. The total area of the lateral margins *𝒜l* is given by 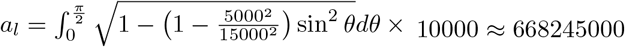 microns squared.

Next we compute the deep margin values, starting with those sampled by proxy. Using Eq. (6), and the fact that *x*^*∗*^ = *x*^*†*^, we find it is

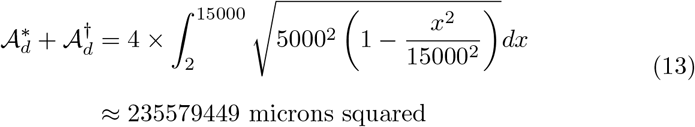

The area of the deep margin is *a*_*d*_ = *π ×* 15000 *×* 5000 *≈* 236000000 microns squared, meaning the percentage *P* of margin sampled by proxy is:

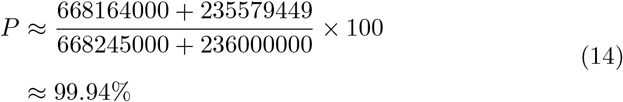

Ironically, we have derived another proof of the “*<* 1%” refrain perpetuated by existing literature. Eq. (14) is less than 100% by about 0.06%, the total area of the margin *directly* sampled by the bread loaf slice, and not accounted for in our calculations. In this example, the whole of the margin is either sampled or sampled by proxy, with the vast majority sampled by proxy.

### 3.2 Example Two: Slices at Ends

Now we assess a hypothetical situation in which a tumor is not cleared until one of the very last sections at each tip. We have arbitrarily selected two mm. from the end as this size is workable for experienced technicians to cut full face sections where smaller pieces may risk being lost in processing or cut at an angle. When we look at a cancer cleared two mm from the end of the specimen with a length of three cm, a width of one cm, and a height of one cm, the percentage of margin assessed is computed as follows. We set *x*^*∗*^ = *x*^*†*^ = 15000*−*2000 = 13000 microns. Using Eq. (1) with the appropriate values, we have

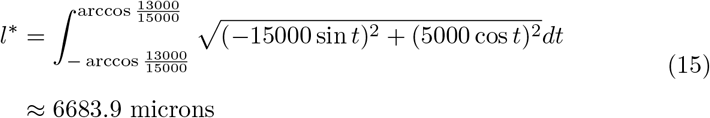

Given that *x*^*∗*^ = *x*^*†*^, we have 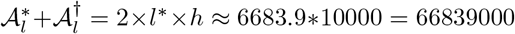 microns squared. The total area of the lateral margin is the same as in the last example, *𝒜*_*l*_ *≈* 668245000 microns squared.

Next we compute the deep margin values, starting with those sampled by proxy. Compared to Eq. (13), Eq. (16) only differs in the range of the integral, replacing 2 microns as the starting point with 13000.

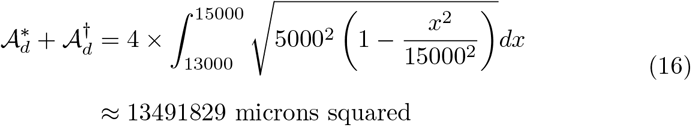

The total area of the deep margin is the same as in the last example, *𝒜*_*d*_ *≈* 236000000 microns squared.

The percentage *P* of margin sampled by proxy is:

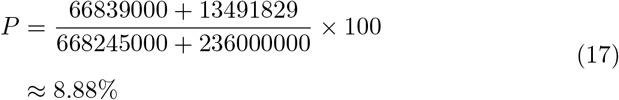

Even in this extreme the percentage of margin sampled by proxy can be seen to be bigger than 1%.

## 4 Discussion

The high cure rates of approximately 95% for basal cell carcinoma managed by simple excision followed by standard processing indicates that this technique is valuable[8]. Reviewing the literature on this subject, we suspect that some assumptions may have inadvertently led to the conclusion that bread loaf sectioning can only give information about 1% or less of the margin. We believe it is natural to focus on the margin of an excisional specimen closest to the center as this margin would seem to be most likely to be involved with the tumor. This is because excisional specimens are intentionally centered around visible cancer. We acknowledge that our assumption that a slice being clear indicates further slices are also clear may not hold true with all kinds of skin cancer. Our assumptions are most pertinent and accurate with tumors that grow as a contiguous, spherical mass with little infiltrative component. This is the case with commonly encountered tumors such as squamous cell carcinoma of keratoacanthoma or nodular basal cell carcinoma. Our model is not as accurate for infiltrative basal cell or squamous cell carcinoma.

Tumor growth in the direction of the minor axis and to the closest margin would intuitively be expected to yield the highest chance of unclear margins, suggesting that clear slices near the tips of the excision are potentially less informative. This suggests that a more meaningful metric could be computed, one which weights a clear portion of the margin by its distance from the center, with close sections receiving more weight than far sections. An appropriate weighting scheme is not immediately apparent, and could even be specific to certain cancer types and their growth patterns, so we leave such endeavors to future work.

The impact of taking additional sections in areas where a tumor approaches the margin also becomes clear. The effectiveness of bread loaf sectioning depends upon the willingness of the pathologist to pursue findings suggestive of tumor extension close to the true surgical margin. Our goal is to point out that bread loaf sectioning can be associated with a broad range of margin analysis, directly and by proxy. If sections are taken every 2-3 mm, the likelihood of tumor extending undetected to a deeper plane between sections is less than if sections are taken only in 10 mm increments along the long axis. Similarly, micronodular, morpheic, infiltrative, or mixed patterns of basal cell carcinomas are all more likely to have finger-like extensions that could escape detection between sections, and this is reflected in the higher rates of margin involvement for these histologic patterns [17]. Because different laboratories have different protocols and procedures, clinicians may not be aware of the method used by their pathologist or dermatopathologist. In fact, it may take months or years before a clinician notices an increased number of skin cancer recurrences, and even then, they may not correlate the change to the way specimens are processed. When we look at the indications for Mohs surgery, many of those indications represent situations where our assumptions (about a tumor free plane indicating that the distal tissue is clear) could be challenged such as with morpheic and infiltrative basal cell carcinomas that may have unusual and unpredictable finger like projections. On the other hand, many tumors encountered in day to day practice are more circumscribed and information about tumor free planes can be inferred with confidence. We believe that it is misleading to conclude that ¡ 0.1% of margins are evaluated when discussing excisions of small, well defined, and low risk basal cell carcinomas [18, 19]. Dermatologists receive significant training in both dermatopathology and dermatologic surgery. The unique skill sets of dermatologists allow for judicious selection of therapeutic options for individual situations. Knowing how tissue is processed in a preferred laboratory and understanding the biologic behavior of various tumor sub-types, should all be part of the decision making process.

We believe that re-considering assumptions and critically evaluating accepted practices help move our specialty forward. Physicians with diverse backgrounds and skill sets may have unique perspectives on common problems. When assumptions are made and reported in peer reviewed journals, they may be perpetuated for long periods of time through “citogenesis” [20, 21, 22]. A fresh look may provide the opportunity to reassess and better understand everyday treatments in a new context. We affirm that the complete circumferential and deep margin assessment accomplished by Mohs surgery has the highest cure rate and is the treatment of choice for histologically aggressive, recurrent, or tumors in areas where tissue conservation is critical. The complete circumferential peripheral and deep margin assessment of Mohs surgery is associated with the highest cure rates and is currently the best method available for full margin evaluation.

In conclusion, we argue that the bread loaf technique can be associated with variable margin analysis but is not limited to ¡ 0.1% of the margin. Bread loaf sections offer significant and valuable margin assessment, especially for less aggressive tumors such as basal cell carcinomas of nodular or superficial type. The willingness of the pathologist to take frequent sections at close intervals, request additional sections when tumor extends close to the margin, and ensure that technicians cut sections full face without folds or gaps all impact results. Currently there is no regulation on the width of slices that different labs take when analyzing a specimen. Nonetheless, in many situations, simple excision followed by bread loaf sectioning remains a very useful tool in the dermatologic armamentarium. Clinicians that understand the risks and benefits of specific techniques available to them are in the best position to make educated decisions that provide good value for their patients.

## Data Availability

Not applicable.

